# Comparable Neutralization of the SARS-CoV-2 Omicron BA.1 and BA.2 Variants

**DOI:** 10.1101/2022.02.06.22270533

**Authors:** Jingyou Yu, Ai-ris Y. Collier, Marjorie Rowe, Fatima Mardas, John D. Ventura, Huahua Wan, Jessica Miller, Olivia Powers, Benjamin Chung, Mazuba Siamatu, Nicole P. Hachmann, Nehalee Surve, Felix Nampanya, Abishek Chandrashekar, Dan H. Barouch

## Abstract

The SARS-CoV-2 Omicron variant (B.1.1.529) has three major lineages BA.1, BA.2, and BA.3^1^. BA.1 rapidly became dominant and has demonstrated substantial escape from neutralizing antibodies (NAbs) induced by vaccination^2-4^. BA.2 has recently increased in frequency in multiple regions of the world, suggesting that BA.2 has a selective advantage over BA.1. BA.1 and BA.2 share multiple common mutations, but both also have unique mutations^1^ (**Fig. 1A**). The ability of BA.2 to evade NAbs induced by vaccination or infection has not yet been reported. We evaluated WA1/2020, Omicron BA.1, and BA.2 NAbs in 24 individuals who were vaccinated and boosted with the mRNA BNT162b2 vaccine^5^ and in 8 individuals who were infected with SARS-CoV-2 (**Table S1**).

---

Following the initial two immunizations with BNT162b2, median pseudovirus NAb responses were 658, 29, and 24 to WA1/2020, Omicron BA.1, and BA.2, respectively (**Fig. 1B**). These data demonstrate a 23-fold and 27-fold reduction of median NAb titers to BA.1 and BA.2 compared with WA1/2020. Six months following initial vaccination, median NAb responses declined to 129, <20, and <20 to WA1/2020, Omicron BA.1, and BA.2, respectively. Two weeks following the third boost with BNT162b2, median NAb responses increased substantially to 6,539, 1,066, and 776 to WA1/2020, Omicron BA.1, and BA.2, respectively, reflecting a 6.1-fold and 8.4-fold reduction of median NAb titers to BA.1 and BA.2 compared with WA1/2020 (**Fig. 1B**). Median BA.2 NAb titers were 1.4-fold lower than BA.1 NAb titers.

**Figure 1.**
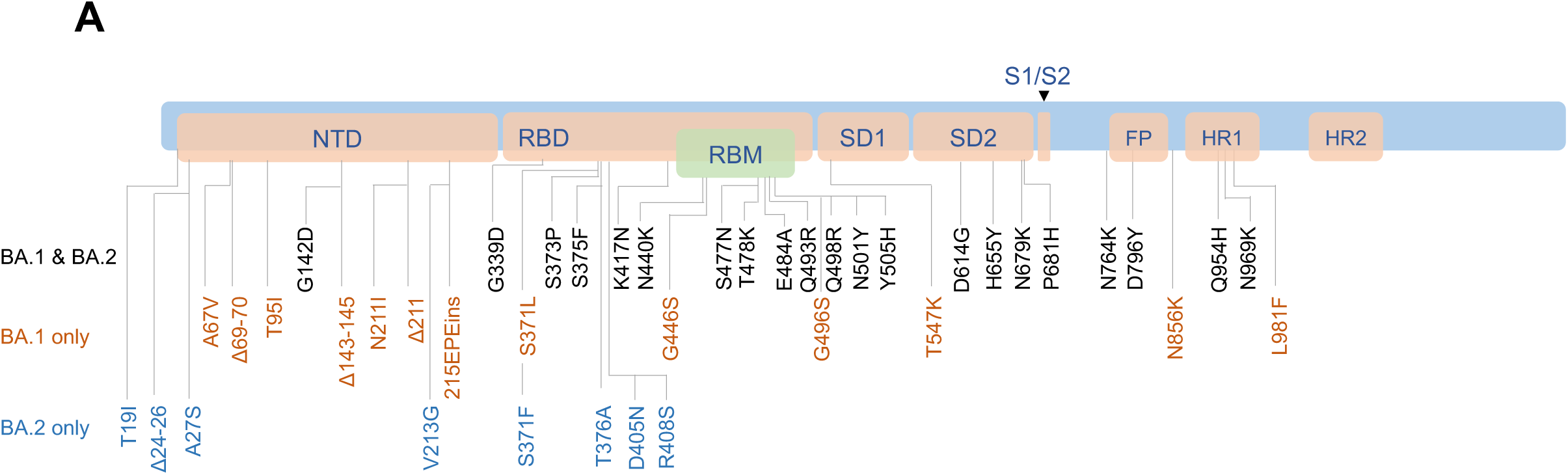
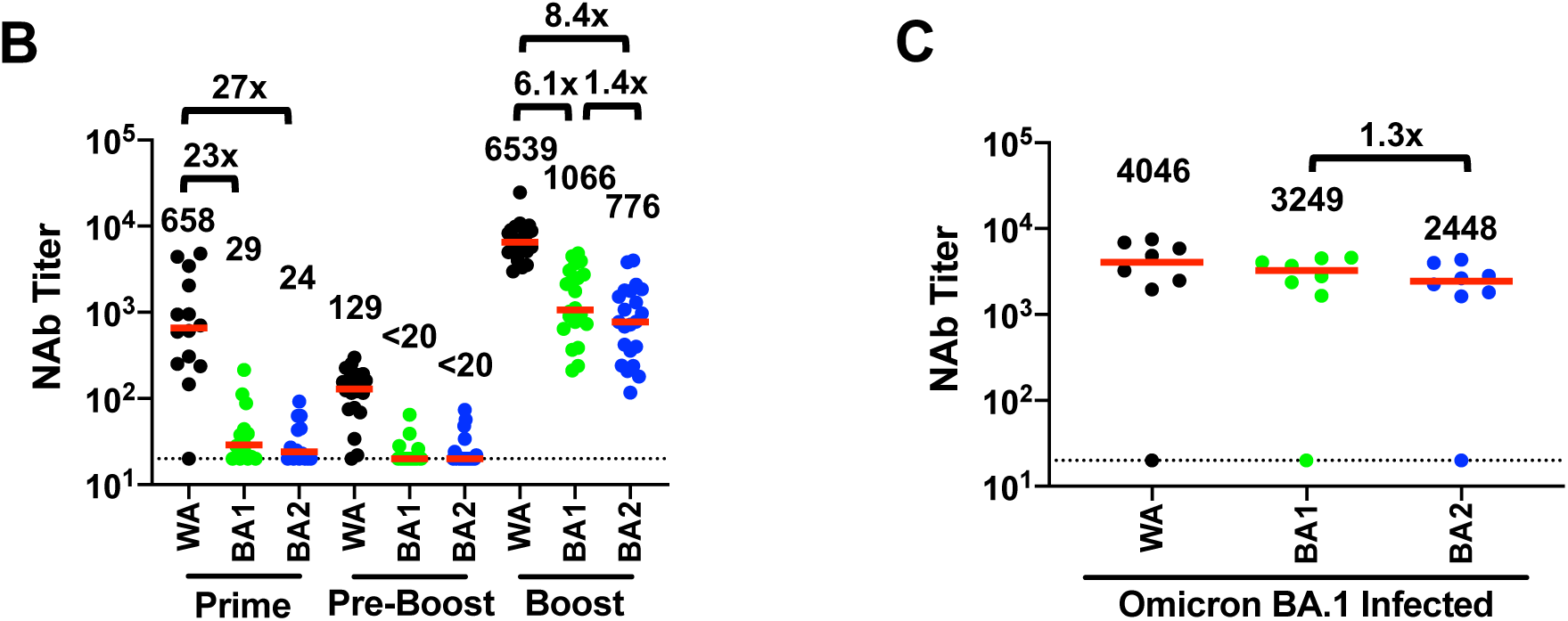
Neutralizing antibody responses to Omicron BA.1 and BA.2. **A**. Cartoon showing BA.1 and BA.2 mutations in the SARS-CoV-2 Spike. NTD, N-terminal domain; RBD, receptor binding domain; RBM, receptor binding motif; SD1, subdomain 1; SD2, subdomain 2; FP, fusion peptide; HR1, heptad repeat 1; HR2, heptad repeat 2. **B**. Neutralizing antibody (NAb) titers by a luciferase-based pseudovirus neutralization assay in individuals two weeks following initial BNT162b2 vaccination (Prime), prior to boost (Pre-Boost), and two weeks following the third boost with BNT162b2 (Boost). **C**. NAb titers in 8 individuals following infection with SARS-CoV-2 Omicron BA.1, of whom 7 were vaccinated. The individual with negative NAb titers was unvaccinated and was sampled 4 days following diagnosis and hospitalization with severe COVID-19 pneumonia. Responses were measured against the SARS-CoV-2 WA1/2020, Omicron BA.1, and BA.2 variants. Medians (red bars) are depicted and shown numerically with fold differences.

We next evaluated NAb titers in 8 individuals (**Table S1**) who were a median of 14 days after diagnosis of SARS-CoV-2 infection, during a time when >99% of new infections were Omicron BA.1. Median NAb responses were 4,046, 3,249, and 2,448 to WA1/2020, Omicron BA.1, and BA.2, respectively (**Fig. 1C**), demonstrating high NAb titers to BA.1 and 1.3-fold lower NAb titers to BA.2. The individual with negative NAb titers was unvaccinated and was sampled 4 days following diagnosis.

These data demonstrate that NAb titers to BA.2 were overall similar to BA.1 but trended 1.3-1.4 fold lower. A third boost with BNT162b2 was required for induction of consistent NAb titers to BA.2, similar to BA.1^3,4^. Moreover, vaccinated individuals infected with BA.1 developed robust NAb titers to BA.2, indicating a substantial degree of cross-reactive natural immunity. These findings have important public health implications and suggest that the increasing BA.2 frequency in the context of the BA.1 surge is likely related to increased transmissibility rather than enhanced immunologic escape.

## Data Availability

All data produced in the present work are contained in the manuscript

## Data sharing

J.Y., A.Y.C., and D.H.B. had full access to all the data in the study and take responsibility for the integrity of the data and the accuracy of the data analysis. All data are available in the manuscript or the supplementary material. Correspondence and requests for materials should be addressed to D.H.B.

## Author Contributions

J.Y., A.Y.C., D.H.B.: conceptualization, formal analysis, resources, investigation, data curation, writing-original draft, writing-review & editing, visualization, supervision J.D.V., H.W., J.M., O.P., B.C., M.S., N.H., N.S., F.N., A.C., M.R.: investigation, methodology, data curation, writing-review & editing

## Acknowledgements

The authors thank the study participants, the Center for Virology and Vaccine Research, and the Department of Obstetrics and Gynecology for enrollment, collection, and processing samples for the BIDMC COVID-19 Biorepository.

## Supplementary Methods

### Study population

A specimen biorepository at Beth Israel Deaconess Medical Center (BIDMC) obtained samples from individuals who received a SARS-CoV-2 vaccine and/or had a history of SARS-CoV-2 infection. The BIDMC institutional review board approved this study (2020P000361). All participants provided informed consent. For the group of individuals receiving three doses of BNT162b2, participants were excluded if they had a history of SARS-CoV-2 infection, received other COVID-19 vaccines, or received immunosuppressive medications

### Pseudovirus neutralizing antibody assay

The SARS-CoV-2 pseudoviruses expressing a luciferase reporter gene were used to measure pseudovirus neutralizing antibodies. In brief, the packaging construct psPAX2 (AIDS Resource and Reagent Program), luciferase reporter plasmid pLenti-CMV Puro-Luc (Addgene) and spike protein expressing pcDNA3.1-SARS-CoV-2 SΔCT were co-transfected into HEK293T cells (ATCC CRL_3216) with lipofectamine 2000 (ThermoFisher Scientific). Pseudoviruses of SARS-CoV-2 variants were generated by using WA1/2020 strain (Wuhan/WIV04/2019, GISAID accession ID: EPI_ISL_402124), Omicron B.1.1.529 BA.1 (GISAID ID: EPI_ISL_7358094.2), or BA.2 (GSAID ID: EPI_ISL_6795834.2). The supernatants containing the pseudotype viruses were collected 48h after transfection; pseudotype viruses were purified by filtration with 0.45-μm filter. To determine the neutralization activity of human serum, HEK293T-hACE2 cells were seeded in 96-well tissue culture plates at a density of 2 × 10^4^ cells per well overnight. Three-fold serial dilutions of heat-inactivated serum samples were prepared and mixed with 50 μl of pseudovirus. The mixture was incubated at 37 °C for 1 h before adding to HEK293T-hACE2 cells. After 48 h, cells were lysed in Steady-Glo Luciferase Assay (Promega) according to the manufacturer’s instructions. SARS-CoV-2 neutralization titers were defined as the sample dilution at which a 50% reduction (NT50) in relative light units was observed relative to the average of the virus control wells.

**Table S1.** Study population.

|  | BNT162b2 (3 dose)<br>N=24 | COVID-19 Infection<br>N=8 |
| --- | --- | --- |
| **Age (years), median (range)** | 34 (23-69) | 37 (23-60) |
| **Sex at birth, female** | 21 (88) | 6 (75) |
| **Race** |  |  |
| White | 20 (83) | 6 (75) |
| Asian | 2 (8) | 0 |
| Black | 1 (4) | 0 |
| More than one race | 1 (4) | 0 |
| Unknown/Other* | 0 | 2 (25) |
| **Ethnicity** |  |  |
| Hispanic or Latino | 2 (8) | 1 (13) |
| Non-Hispanic | 22 (88) | 6 (75) |
| Other | 1 (4) | 1 (13) |
| **Medical condition** |  |  |
| Obese (BMI $\geq 30$ kg/m<sup>2</sup>) | 1 (4) | 2 (25) |
| Hypertension | 2 (8) | 2 (25) |
| Diabetes | 2 (8) | 3 (50) |
| Immunosuppression | 0 | 1 (13) |
| Pregnant | 1 (4) | 3 (50) |
| **COVID-19 vaccination history** |  |  |
| BNT162b2 (3 doses) | 24 (100) | 2 (25) |
| mRNA-1273 (3 doses) | 0 | 2 (25) |
| BNT162b2 (2 doses) | 0 | 2 (25) |
| Ad26.COV2.S (1 dose)/mRNA-1273 (1 dose) | 0 | 1 (13) |
| Unvaccinated | 0 | 1 (13) |
| **COVID-19 disease severity (NIH scale)** |  |  |
| Asymptomatic | N/A | 1 (13) |
| Mild | N/A | 5 (63) |
| Moderate | N/A | 0 |
| Severe | N/A | 2 (25) |
| Critical | N/A | 0 |
| **Days from primary vaccine series to booster vaccine\*\*, median (IQR)** | 254 (248-258) | 268 (244-295); n=5 |
| **Days from last vaccine dose to positive PCR test, median (IQR)** | N/A | 113 (27-219) |
| **Days from positive PCR test to sampling, median (IQR)** | N/A | 14 (5-22) |

Data displayed as median (range or interquartile range, IQR) and n (%). BMI, body mass index; PCR, polymerase chain reaction for SARS-CoV-2; pregnant designation reflects time of sampling. \*Self-identified race as Brazilian and Colombian. \*\*Time from 2<sup>nd</sup> mRNA vaccine to 3<sup>rd</sup> vaccine or time from single Ad26.COV2.S vaccine to 2<sup>nd</sup> vaccine.

## References

1. Viana R, Moyo S, Amoako DG, et al. Rapid epidemic expansion of the SARS-CoV-2 Omicron variant in southern Africa. Nature 2022.

2. Cele S, Jackson L, Khoury DS, et al. Omicron extensively but incompletely escapes Pfizer BNT162b2 neutralization. Nature 2021.

3. Schmidt F, Muecksch F, Weisblum Y, et al. Plasma Neutralization of the SARS-CoV-2 Omicron Variant. N Engl J Med 2021.

4. Liu L, Iketani S, Guo Y, et al. Striking Antibody Evasion Manifested by the Omicron Variant of SARS-CoV-2. Nature 2021.

5. Polack FP, Thomas SJ, Kitchin N, et al. Safety and Efficacy of the BNT162b2 mRNA Covid-19 Vaccine. N Engl J Med 2020.

